# Cognitive Ageing in Healthy Adults in India: A Cross-Sectional Multimodal Study Protocol

**DOI:** 10.64898/2026.09.04.26362232

**Authors:** Hurshitha Vasudevan, Vidya Chelluri, Govindhaswamy Umapathy, PK Vinod, Naresh Babu V. Sepuri, Thanuja Krishnamoorthy, S. Bapi Raju, Bhaktee Dongaonkar

## Abstract

**Introduction:** The population of older adults in India is projected to exceed 230 million within the next decade. With increasing life expectancy, maintaining cognitive and neurobiological health becomes essential for ensuring a good quality of life. This study is a comprehensive multimodal investigation of aging and cognition in a healthy Indian cohort. The aim is to characterize cognitive, neurobiological, physiological, psychological, and molecular trajectories and their associations across the adult lifespan in India.

**Methods and Analysis:** Data will be collected cross-sectionally from healthy young (19–35 years, *n* = 100), middle-aged (40–55 years, *n* = 100), and older adults (60–75 years, *n* = 100) with regional nativity and residence in Telangana or Andhra Pradesh. Individuals with cognitive impairment or any chronic health condition will be excluded. Assessments will include standardized cognitive tests, mental health questionnaires, brain imaging, heart rate variability, and serum samples for standard blood tests and epigenetic analyses. Hair cortisol and serum inflammatory markers will also be collected to profile stress and systemic inflammation, respectively. Data from this study will help to profile healthy aging in an Indian cohort, thereby addressing critical gaps in clinical research and health policy.

**Ethics and Dissemination:** This study was approved by the Institutional Review Board at IIIT Hyderabad (IIITH-IRB-PRO-2023-11). All participants will give written consent. Results from this study will be presented at conferences and published in peer-reviewed journals. Data will also be curated for an interactive web-portal to enable researchers, clinicians, and policy makers to understand the data in meaningful ways.

**Strengths and Limitations:**

- Most studies in aging focus on risk factors and progression of degenerative conditions. What healthy aging really means is not well understood.
- This study will recruit healthy Indian adults free from any chronic health problem and with regional nativity and residence to minimize geo-climatic effects and genetic heterogeneity.
- Hair cortisol and serum-inflammation levels may provide unique insights in healthy aging.
- The comprehensive data from this study will provide a profile of healthy aging in an Indian cohort that is highly vulnerable to metabolic disorders. It will enable clinicians, researchers, and policy makers to plan and promote healthy aging.
- The restricted native sample population and sample size may not capture the divergent trajectories in aging across other demographic populations within and outside India.
- Though cross-sectional data will not capture the changes that longitudinal data can, it will offer an opportunity to compare psychosocial and physiological measures across different age groups.

## INTRODUCTION

The aging population is growing rapidly worldwide and within India it is projected to reach 230 million by 2036, nearly 15% of the Indian population.^1^ To prepare and cope with the implications of the growing elderly population, the United Nations has declared 2021-2030 as the Decade of Healthy Aging^2^. As lifespan increases, neurocognitive, physiological, and psychosocial data from healthy individuals are needed to characterize normative changes associated with aging.

In the last two decades several ageing studies have been conducted, however, the data is predominantly from the Caucasian population.^3–6^ Translating results to the Indian population has been a challenge for clinicians and mental health professionals in India. The Indian brain is smaller in size.^7–9^ Differences in brain size and surface area or volume of specific regions can manifest in different clinical outcomes.^10–13^ Further, the compensatory neural mechanisms that remain active in older adults^14–15^ may have different trajectories in different populations. Factors like lifestyle and diet also vary across ethnicities which are known to dysregulate metabolic activity, accelerate aging, or trigger epigenetic changes.^16–18^ Thus, population-specific studies on healthy aging are important.

There are significant aging studies underway in India. The Longitudinal Aging Study of India (LASI) has surveyed socio-economic and health factors in ∼72000 individuals > 45 years over 10 years to understand the prevalence of dementia and associated risk factors.^19^ A subset of the LASI participants > 60 years will undergo comprehensive assessments for late-life dementia.^20^ Another systematic study is underway to study biochemical and neurocognitive risk factors of dementia in a rural cohort > 45yrs in age, with follow-ups every 2-3 years.^21^ These studies are focused towards early diagnosis and characterization of neurodegenerative conditions in individuals above 45yrs. Other Indian studies on older adults have reported effects of physical inactivity, education, gender, and alcohol and nicotinic substances on cognitive impairment.^22–26^ Overall, the focus has been to understand factors that result in cognitive impairment or degenerative conditions. A systematic study of healthy aging and cognition in India is lacking.

To study healthy aging and cognition, chronic health conditions that may affect cognition need to be ruled out. India has a high prevalence of metabolic conditions such as hypertension and diabetes, adding up to 68% of chronic diseases in older adults.^23,27^ Chronic metabolic conditions affect cognition.^28–29^ A recent study reported impairment on tests of attention and executive function in community dwelling Indian older adults with high levels of blood sugar.^30^ Thus, screening for metabolic health is also important, especially in the context of healthy aging in India.

In addition, psychosocial stress, mental health, lifestyle, and diet may increase systemic inflammation which may trigger epigenetic changes that accelerate aging and alter cognitive function.^31–32^ Therefore, measures of chronic stress and inflammation need to be incorporated in the context of healthy aging and cognition.

This study was designed to collect multimodal data inclusive of psychological, physiological, socioeconomic, cognitive, neuroimaging, hair and serum samples, to provide a holistic understanding of healthy ageing in an Indian cohort. Data will be collected from three age groups: young, middle-aged, and older adults. To reduce genetic and geo-climatic heterogeneity, this study will include healthy individuals residing in the Telangana or Andhra Pradesh regions of India, with at least three generations of nativity from the same regions.

## RESEARCH AIM

To characterize age-related changes in cognitive, neurobiological, physiological, psychological, and molecular measures in healthy adults across the adult lifespan in India.

## RESEARCH OBJECTIVES

1. To examine the patterns of changes in cognitive, neuroimaging, physiological, psychological, and molecular markers across healthy adult age groups.
2. To examine relationships among cognitive, neuroimaging, physiological, psychological, and molecular measures across healthy adult age groups.
3. To characterize variability in cognitive, neuroimaging, physiological, and molecular ageing among healthy adults.
4. To examine associations among stress, inflammation, and neurocognitive function across healthy adult age groups.
5. To establish preliminary age-specific normative profiles for cognitive, neurobiological, physiological, and biological measures in healthy Indian adults.

## METHODS AND ANALYSIS

### Study Design and Sample Size

This study will recruit 300 healthy community-dwelling participants in three age cohorts: n=100 young adults (19-35 years), n=100 middle-aged adults (40-55 years), and n=100 older adults (60-75 years) with a balanced number of men and women in each group. Sample size estimation was conducted using an a priori power analysis based on moderate effect size (Cohen’s f ∼ 0.25), statistical power of 80%, and α = 0.05 from previous studies for different groups of outcomes. For three age groups using analysis of variance (ANOVA), power calculations indicated that a minimum of 75 participants per age group. To accommodate loss of data from exclusion criteria, we planned for 100 in each group. Data collection began in 2023 and will continue until end of 2026. The protocol is summarized in Figure 1.

**Figure 1:**
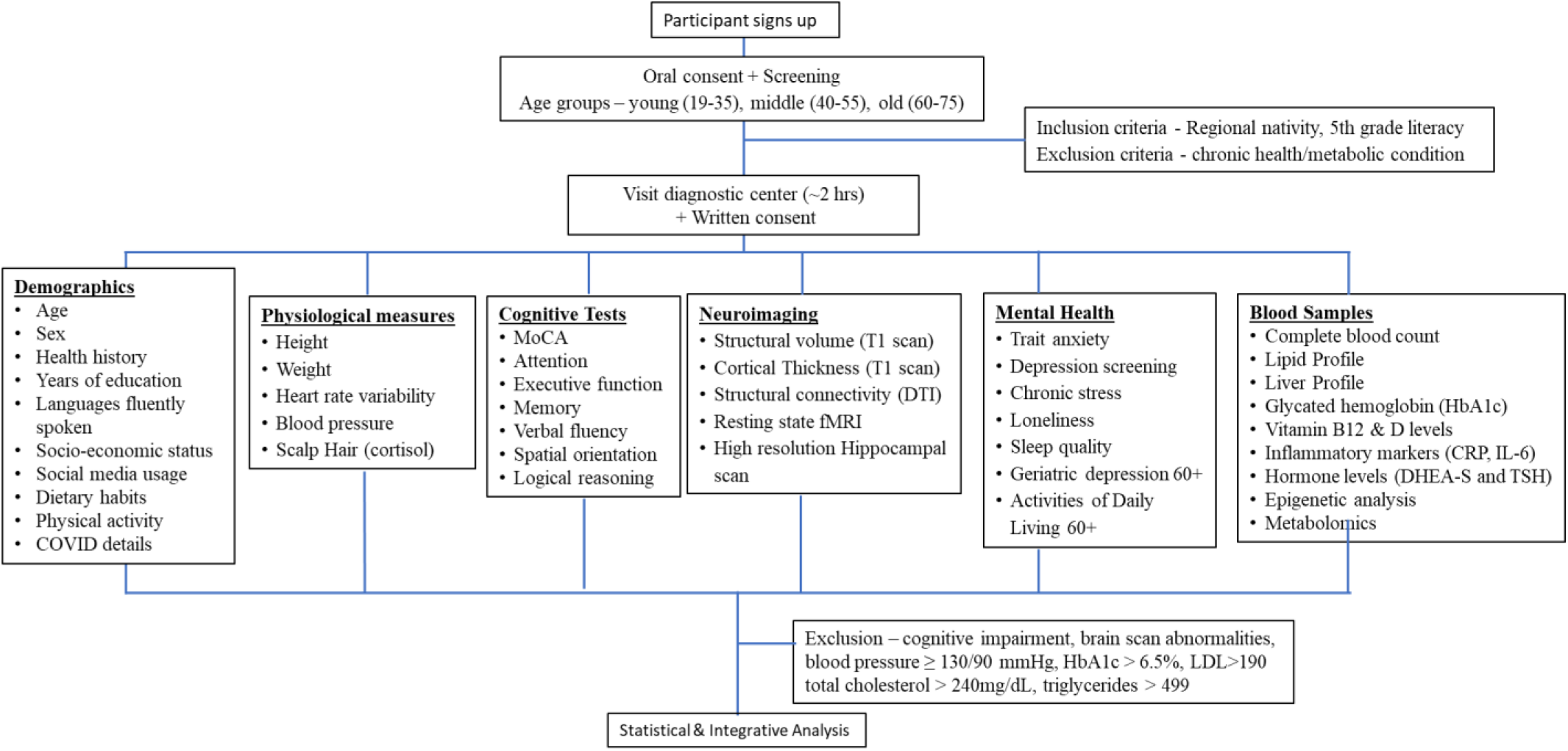
Summary of the protocol and outcome measures.

### Participant Recruitment

Posters about the study will be circulated. Senior well-being centres, rotary clubs, and old age homes will be contacted to bring awareness about this study. Finding healthy older adults can be challenging, therefore snowball referrals will also be encouraged. The study team will reach out to participants from a wide range of socioeconomic strata through word of mouth. Interested participants will directly contact the study team using a phone number on the poster. Oral consent will be obtained over a phone call or in-person to establish eligibility based on the inclusion/exclusion criteria listed below. A visit to the diagnostic centre (Lucid Diagnostics, Hyderabad, India) will then be arranged, where participants will give written consent before proceeding with further testing. Consent forms with the objectives, methodology, and purpose of the study, will be available in English, Telugu, and Hindi.

Imaging data and blood samples will be collected at the diagnostic centre. Cognitive tests and other measures will be collected either at the diagnostic center or at the participant’s home, for convenience.

### Inclusion Criteria

1. Regional nativity (raised and living in Telangana/Andhra Pradesh from 3 generations). This is to reduce geo-climatic and genetic heterogeneity.
2. Basic reading and writing proficiency required, equivalent to a 5th-grade level, to follow instructions to perform cognitive tests.

### Exclusion Criteria

1. Any prescribed medication for more than 3 months.
2. Chronic health conditions (e.g. diabetes, hypertension, asthma, thyroid, etc.)
3. Cognitive impairment, psychiatric conditions, neurological or neurodegenerative conditions, or brain injury.
4. MRI compatibility issues: Presence of metal implants in the body or claustrophobia.
5. Severe hearing or vision difficulties
6. Pregnancy

### Assessments and Data Collection

Demographic data and mental health scales summarized in Table 1.

**Table 1:**
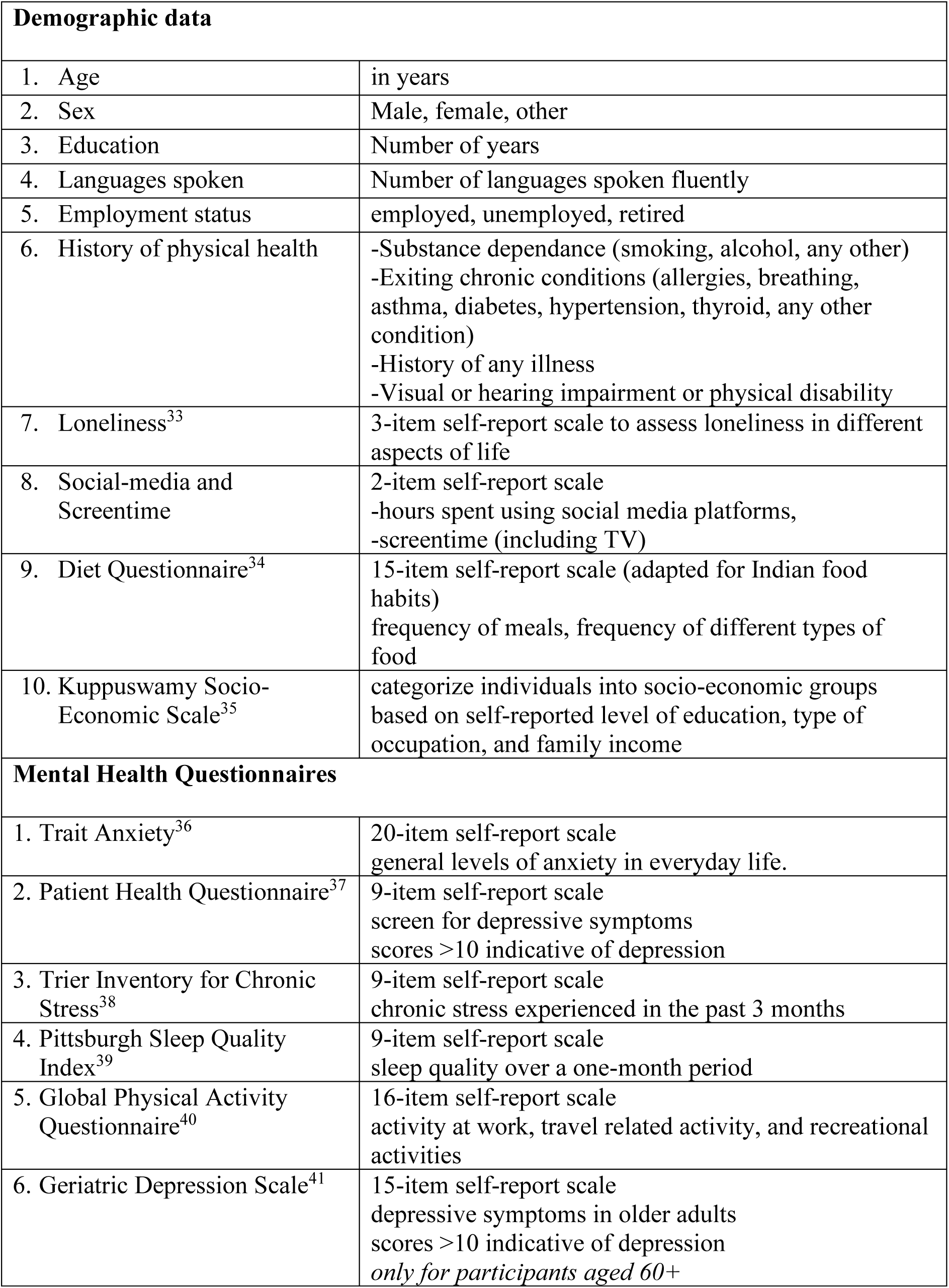
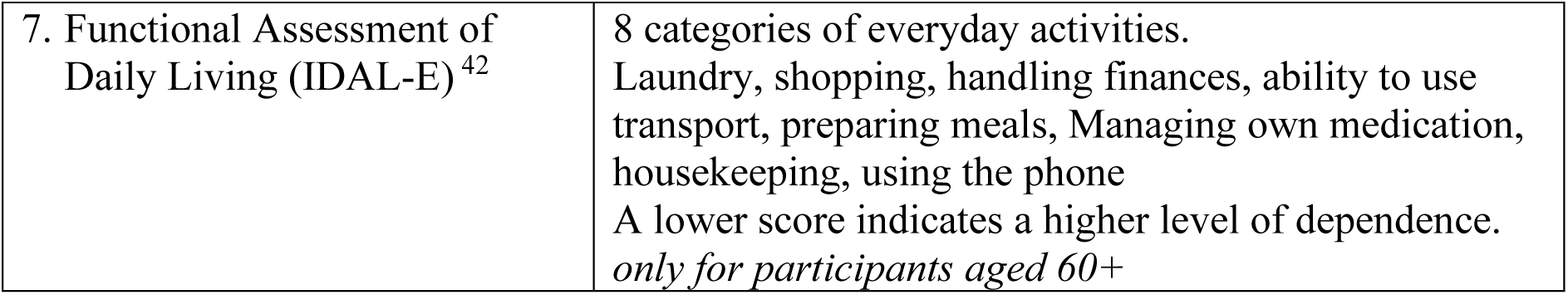
Demographic data and mental health measures.

### Neuroimaging

Multimodal neuroimaging data will be collected in a 3T Siemens Magnetom Lumina MRI scanner with high spatial resolution at the diagnostic centre. Scanning duration for each participant will be approx. 45 mins. The protocols in Table 2 will be used to collect neuroimaging data based on international standards such as the Alzheimer’s Disease Neuroimaging Initiative (ADNI).^32,43–45^

**Table 2:** Standard neuroimaging protocols to be implemented in the study.

| Type of Scan | TR (ms) | TE (ms) | FoV (mm) | Slice Thickness (mm) | Purpose |
| --- | --- | --- | --- | --- | --- |
| 3D T1 weighted | 2200 | 3.43 | 256 | 1.0 | To observe gray and white matter distribution |
| T2* weighted scans | 3200 | 408 | 250 | 1.0 | To rule out any brain-related abnormalities |
| Resting state functional MRI (rs-fMRI) | 3000 | 30 | 192 | 3.5 | To observe brain networks that are active at rest (eyes closed) |
| Diffusion Tensor Imaging (DTI) - 66 directions | 9800 | 104 | 250 | 2.0 | To assess white matter tracts for structural connectivity |
| Accelerated High Resolution of Hippocampus | 5850 | 100 | 220 | 2.0 | To observe minute changes in the hippocampal structure |

All neuroimaging data will be processed using standardized pipelines implemented in well-established Neuroimaging tools. Structural T1- and T2-weighted scans will be pre-processed using the FreeSurfer toolbox,^46^ including skull stripping, bias correction, and segmentation, followed by cortical thickness estimation, gray/white matter density mapping, volumetric analysis, and voxel-based morphometry. Diffusion Tensor Imaging (DTI) data will be processed with MRtrix^47^ for motion and eddy current correction, tensor fitting, tractography, and subsequent network-based structural connectivity analysis to extract diffusion-based metrics. Resting-state fMRI will be pre-processed using fMRIPrep,^48^ incorporating motion correction, normalization, and nuisance regression, and subsequently analysed using seed-based functional connectivity, independent component analysis, and graph-theory based approaches. High-resolution hippocampal scans will be processed in FreeSurfer for segmentation and volumetric analysis of hippocampal subfields. Finally, multimodal integration will be performed to model structure–function relationships by linking structural measures (cortical thickness, volumetrics, hippocampal subfields, DTI-based tractography) with functional measures (resting-state networks) to derive comprehensive network-level differences.

### Blood Samples

Approximately 8ml of blood will be collected by a phlebotomist, of which 4ml will be used to conduct standard blood tests listed below. All pathology tests, summarized in Table 3, will be performed at the diagnostic center. These tests will help to rule out unknown health conditions in addition to age-related variations.

**Table 3:**
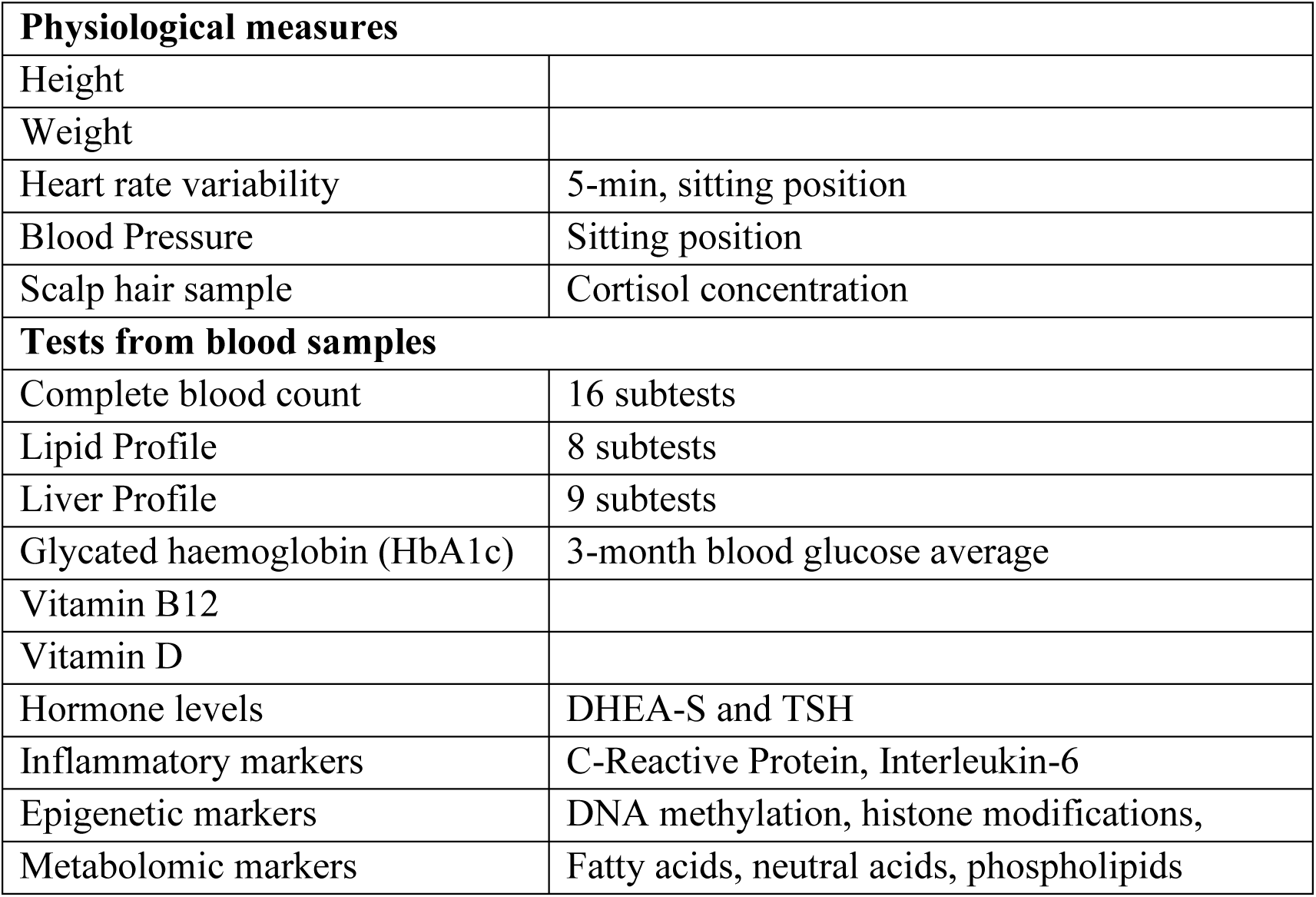
Physiological measures and data from blood samples.

An additional 4 ml of blood will be transported to the Bio-Lab at IIIT-H in an ice box. Within 4 hours, blood components and plasma will be isolated using a standardized gradient centrifugation protocol. Briefly, the blood sample will be diluted with Phosphate-Buffered Saline (PBS) and layered over Histopaque, then centrifuged to separate components. The plasma (top layer) and Peripheral Blood Mononuclear Cells (PBMC)-containing middle layer will be collected into separate tubes. PBMCs will be washed with PBS and split into two aliquots for DNA and RNA isolation. The isolated samples will be stored at –80 °C in the Bio-Lab at IIIT-H. Further analyses will include epigenetic profiling to identify markers of DNA methylation and histone modifications, along with transcriptome analysis for RNA related molecular activity.

The isolated plasma samples will be used for metabolomic and lipidomic analysis which will provide a glimpse into the biochemical activity in response to aging, stress, and lifestyle. The plasma samples will be mixed with methanol and methyl tertiary butyl ether to extract metabolites, followed by phase separation through centrifugation. The organic and aqueous layers will be collected separately and dried. For polar metabolites (glycolysis, lipids), samples will undergo methoxyamine treatment and silylation to form stable, volatile derivatives for gas chromatography-mass spectrometry (GC-MS). For lipidomics, fatty acids will be converted to fatty acid methyl ester using methanolic HCl, extracted with hexane, washed with brine, dried with sodium sulfate, and evaporated. The residue will be reconstituted in hexane and the prepared samples will be injected into GC-MS for metabolite identification using standards.

### Hair Sample for Cortisol

Cortisol, the stress hormone, accumulates in the hair follicles^49^. Every inch away from the hair follicle carries cortisol deposited in the past 2-3 weeks, approximately. Considering the average growth of hair, 3 inches corresponds to approximately the past 3-4 months of a participant’s life. A bunch of hair strands will be collected from the crown of the participant’s head. These hair strands will be cut at ∼3 inches from the follicle end and will be stored at −20 degrees. Hair cortisol deposited in the hair will be extracted and analysed.

Briefly, hair samples will be thoroughly washed with isopropanol and dried at 40°C for 2-3 days and pulverized into fine powder. About 10 mg of hair powder will be incubated overnight in methanol at 52°C with gentle shaking, then centrifuged. The methanol extract will be collected and evaporated under nitrogen. The dried residue will be reconstituted in PBS (pH 8.0), vortexed, and stored at –20°C until further analysis^50–51^. Cortisol concentrations will be determined using a standard cortisol enzyme immunoassay method^51^.

### Physiological Measures

Height, weight, blood pressure, and pulse rate will be collected for every participant. Heart rate variability (HRV) will be recorded for 5 minutes in a sitting position using a Scosche-Rhythm armband and will be analyzed in time and frequency domains. HRV metrics indicate the proportion of sympathetic vs parasympathetic activity in the nervous system^52^.

### Cognitive Tests

Standard cognitive tests will comprehensively assess different domains of cognitive functions which are crucial for the everyday functioning of a healthy individual. Cognitive tests included in the study are summarized in Table 4. Most tests will be conducted in paper-and-pencil format, while a few will be administered on a laptop.

**Table 4.** Cognitive tests and scoring outcomes.

| **Cognitive Domain** | **Cognitive Test** | **Cognitive Ability Assessed** | **Scoring Format** |
| --- | --- | --- | --- |
| Multi-domain | Montreal Cognitive Assessment (MoCA)<sup>53</sup> | Screening for Mild Cognitive Impairment | Score out of 30 |
| Attention & Executive Function | Trail Making Test<sup>54</sup> | Visuo-motor coordination, working memory, attention switching | Time taken (secs) |
|  | Sustained Attention to Response Test (SART)<sup>55</sup> | Response inhibition, sustained attention | Accuracy score + Reaction time (secs) |
|  | Digit Cancellation Test<sup>56</sup> | Processing speed | Total digits cancelled in 90secs |
|  | N-Back<sup>57</sup> | Working memory | Accuracy score + Reaction time (secs) |
| Memory Processes | Rey's Auditory Verbal Learning Test (RAVLT)<sup>58</sup> | Verbal memory | Rate of learning, delayed recall, interference, recognition |
|  | Weschler's Visual Reproduction<sup>59</sup> | Visuo-spatial memory | Delayed recall |
|  | Optimized-Mnemonic Separation Task (O-MST)<sup>60</sup> | Pattern separation (hippocampal dependent) | Recognition memory score |
| Spatial Orientation | Clock Test Activity<sup>61</sup> | Allocentric & egocentric spatial processing | Accuracy score |
| Language | Category Fluency<sup>54</sup> | Semantic fluency | No. of words |
|  | Letter Fluency<sup>54</sup> | Phonemic fluency | No. of words |
| Reasoning | Raven's Progressive Matrices<sup>62</sup> | Logical reasoning | Accuracy score |

### Planned Statistical Analysis

Statistical analyses for such multimodal data will include descriptive statistics and age-group comparisons using General Linear Models (GLM), Analysis of Covariance (ANCOVA), and regression analyses, with relevant covariates. Age will be examined both categorically and continuously, with nonlinear trajectories explored where appropriate. Region-of-interest (ROI)–based mapping of brain regions and their associated cognitive functions will be performed to determine whether age-related differences in volume, cortical thickness, and intra-regional versus inter-regional connectivity are associated with corresponding cognitive changes across healthy age groups. Associations among stress, inflammation, cognition, and health measures will be assessed using correlation, multivariable regression, and exploratory mediation analyses. Multimodal relationships will be examined using Principal Component Analysis (PCA), Partial Least Squares (PLS), or Canonical Correlation Analysis (CCA), while clustering or normative modelling may be used to characterize individual variability in healthy ageing.

### Integrative Omics Analysis

The comprehensive nature of this dataset will enable integration across molecular, neurobiological, and physiological domains of healthy aging. Blood-based multi-omics data (epigenomics, transcriptomics, and metabolomics) will be integrated with neurocognitive measures, brain imaging features, immune and inflammatory profiles, and metabolic markers to derive a multidimensional view of healthy aging. Differentially methylated regions will be linked to corresponding transcriptional changes and mapped onto biological pathways across age groups. Gene regulatory network analysis will further identify key transcription factors involved in these processes. Associations between epigenetic and transcriptional changes and metabolomic profiles will provide mechanistic insights into how molecular alterations translate into functional changes.

Unsupervised clustering of omics features may reveal molecularly distinct subgroups within the healthy cohort. Such subgroups could represent different aging trajectories—for instance, individuals exhibiting younger molecular profiles relative to their chronological age versus those showing early molecular signatures of aging. Supervised regression models will be trained on molecular and/or multimodal features to predict chronological age, with residuals representing the difference between predicted and actual age, thereby providing a quantitative estimate of biological aging. Overall, the dataset will support the development of predictive models to identify deviations from healthy aging based on multimodal features.

### Data Privacy and Storage

Strict protocols will be followed for data privacy and storage. Each participant will be given a study code, and beyond the consent form, there will be no identifiable information that reveals participant identity. Hard copies of consent forms and cognitive tests will be stored securely at IIIT-H, with access limited only to authorised personnel. Data from cognitive tests, questionnaires, blood tests, and imaging data will be stored on secured devices. Isolated blood samples will be stored in secured freezers at the Bio-Lab at IIIT-H. The anonymized and analysed data will be stored in secured servers.

### ETHICS AND DESSEMINATION

This study was approved by the Institutional Ethics Committee at IIIT-Hyderabad (IIITH-IRB-PRO-2023-11). All participation will be free of cost and participants will be given a copy of their blood reports and brain scans. In rare cases of abnormal blood report or brain scan, the participant will be advised to visit a clinician.

Data from this study will first be presented at conferences and published in peer-reviewed journals. Data will then be curated to be uploaded to an interactive web-portal where anyone can get insights into the data, including researchers, policy makers, and citizens. Raw data will only be made available to researchers with confidentiality clauses. Data will remain secured in servers located at IIIT Hyderabad.

## DISCUSSION

This study aims to systematically investigate healthy aging by integrating cognitive, neuroimaging, physiological, blood, epigenetic, psychological, and socioeconomic data from participants with a relatively homogeneous background. Data from healthy adults free from chronic health conditions will provide an opportunity to characterize age-related patterns of healthy aging while minimizing the influence of disease-related decline. This is especially crucial in a demographic context like India, where rapidly aging populations are juxtaposed with a rising prevalence of non-communicable diseases such as diabetes and hypertension.

Most current cognitive and physiological aging norms are derived from Western populations, whose life experiences, genetic backgrounds, and health trajectories differ significantly from the Indian population. This study will facilitate clinicians, researchers, and policymakers to plan better assessments and informed interventions by providing population-relevant reference profiles of healthy aging.

A key aspect of this study is its emphasis on a relatively homogeneous participant population. By limiting recruitment to individuals with three generations of regional ancestry and residence (Telangana and Andhra Pradesh), the study attempts to minimize some sources of genetic and environmental variability, thereby facilitating the investigation of subtle epigenetic changes and brain-behaviour correlations in healthy aging.

Comparing young, middle-aged, and older adults will help to characterise age-related differences in cognitive and neural function across the adult lifespan and potentially identify cognitive abilities and neural function that may be preserved in healthy aging. In addition, hair cortisol, inflammatory, metabolic, and epigenetic measures will enable examination of biological processes associated with aging in the absence of chronic disease, particularly in an Indian cohort. Further, the inclusion of sleep, anxiety, HRV, loneliness, and other psychosocial data will provide an opportunity to explore their interaction in the context of healthy aging and whether these factors correlate with cognitive function and brain connectivity across adulthood.

## Data Availability

This manuscript is a study protocol. Does not present, analyze or describe any data.

## ACKNOWLEDGEMENT

We are grateful to Dr. Ramesh Yelagandula for providing support for the epigenetic and transcriptomic analysis. We thank Dr. Gopal Gopala for his unwavering support for this project. We also thank Dr. Dipti Misra for helping with thorough translations of consent forms in regional languages.

## AUTHOR CONTRIBUTIONS

Conceptualization - SBR, BD

Funding Acquisition - BD, SBR

Data Collection - HV, SC

Data Curation - HV, SC, BD

Methodology - HV, SC, BD, GU, NBVS, TK

Statistical Planning - HV, SC, BD, GU, PKV

Integrative Analysis - PKV, BD, SBR

Writing original draft - HV, SC, BD

Reviewing and Editing - HV, SC, BD, GU, PKV, SBR

All authors approved the final version of the manuscript.

## COMPETING INTEREST

We declare no competing or conflict of interest.

## PATIENT CONSENT FOR PUBLICATION

No patients involved. Patient consent not required.

## PATIENT AND PUBLIC INVOLVEMENT

No patients or public are involved in the funding, design, recruitment, analysis or dissemination of this study.

## FUNDING STATEMENT

This work was supported by DST-IHUB-Data at IIIT Hyderabad, grant number (IIITH/IHub/Project/Healthcare/H2-007). IHUB-Data was not involved in the study design nor will it be involved in data collection, analyses, writing, or decisions related to publications.

## Notes

### Competing Interest Statement

The authors have declared no competing interest.

### Author Declarations

This study was approved by the Institutional Ethics Committee at IIIT-Hyderabad (IIITH-IRB-PRO-2023-11). This study will collect data from human subjects for a cognitive and public health study.

